# Administrative Data–Based Staging of the Heart Failure Continuum in a Nationwide Population

**DOI:** 10.64898/2026.07.10.26357788

**Authors:** Jiri Jarkovsky, Jiri Parenica, Klara Benesova, Ales Linhart, Jan Krejci, Filip Malek, Radek Pudil, Petr Ostadal, Jan Belohlavek, Anna Chaloupka, Tomas Palecek, Milos Kubanek, Josef Kautzner, Jiri Hlasensky, Ladislav Dusek, Vojtech Melenovsky, Peter Wohlfahrt

**Affiliations:** Institute of Health Information and Statistics, Prague, Czechia; Department of Internal Medicine and Cardiology, University Hospital Brno, Faculty of Medicine, Masaryk University, Brno, Czechia; 2nd Department of Medicine - Department of Cardiovascular Medicine, First Faculty of Medicine, Charles University and General University Hospital, Prague, Czechia; First Faculty of Medicine, Charles University, Prague, Czechia; First Department of Internal Medicine – Cardioangiology, St. Anne’s University Hospital, Faculty of Medicine, Masaryk University, Brno, Czechia; Department of Cardiology, Na Homolce Hospital, Prague, Czechia; 1st Department of Internal Medicine-Cardioangiology, Faculty of Medicine in Hradec Kralove, Charles University, Hradec Kralove, Czechia; Department of Cardiology, Second Faculty of Medicine, Charles University and Motol University Hospital, Prague, Czechia; Department of Cardiology, Institute for Clinical and Experimental Medicine, Prague, Czechia

**Keywords:** heart failure, staging, prevalence, administrative data, mortality, epidemiology

## Abstract

**Background:** Population-level data on preclinical heart failure (HF) remain limited because most epidemiological studies focus on symptomatic HF. We therefore developed an administrative-data algorithm to classify HF stages across the national population and describe temporal trends, stage transitions, and mortality across the HF continuum.

**Methods:** Using a claims-based staging framework adapted from the Universal Definition of HF, we classified HF stages from ICD-10 codes, prescription records, and medical procedures. We applied this algorithm to the Czech population, linking the National Registry of Reimbursed Health Services to National mortality records from 2015 to 2024.

**Results:** In 2024, 27.8% of the Czech population met criteria for Stage A HF and 8.2% for Stage B. Over 10 years, the prevalence of both preclinical stages increased beyond what could be explained by population aging alone, with age-standardized prevalence rising by 9.5% for Stage A and 19.2% for Stage B. Age-standardized 1-year mortality showed a steep stepwise gradient, from 0.69% in Stage A to 1.69% in Stage B, 3.06% in Stage C, and 7.27% in Stage D. Among 52,172 individuals with incident clinical HF in 2024, more than 95% had previously met administrative criteria for Stage A or Stage B.

**Conclusion:** Administrative surveillance of the HF continuum using routinely collected healthcare data provides a scalable administrative framework for population-level monitoring of HF burden. In Czechia, both preclinical and clinical HF burdens increased over time beyond population aging alone, underscoring the need for earlier preventive strategies targeting preclinical disease.

**Clinical Perspective:**

- We developed a nationwide administrative-data algorithm that classifies the full heart failure continuum (Stages A–D) using routinely collected ICD-10 diagnoses, prescription claims, and procedure codes, showing that both preclinical and clinical HF burdens are rising beyond what population aging explains.
- More than 95% of individuals who developed clinical HF had previously met administrative criteria for Stage A or Stage B, suggesting that progression to overt disease is often preceded by an identifiable preclinical phase that may represent an opportunity for prevention.
- This scalable, low-cost surveillance framework enables health systems to identify at-risk populations, track HF trends over time, and target preventive strategies toward preclinical stages, an approach that could be adapted internationally to inform policy and resource allocation.

**Graphical abstract:** 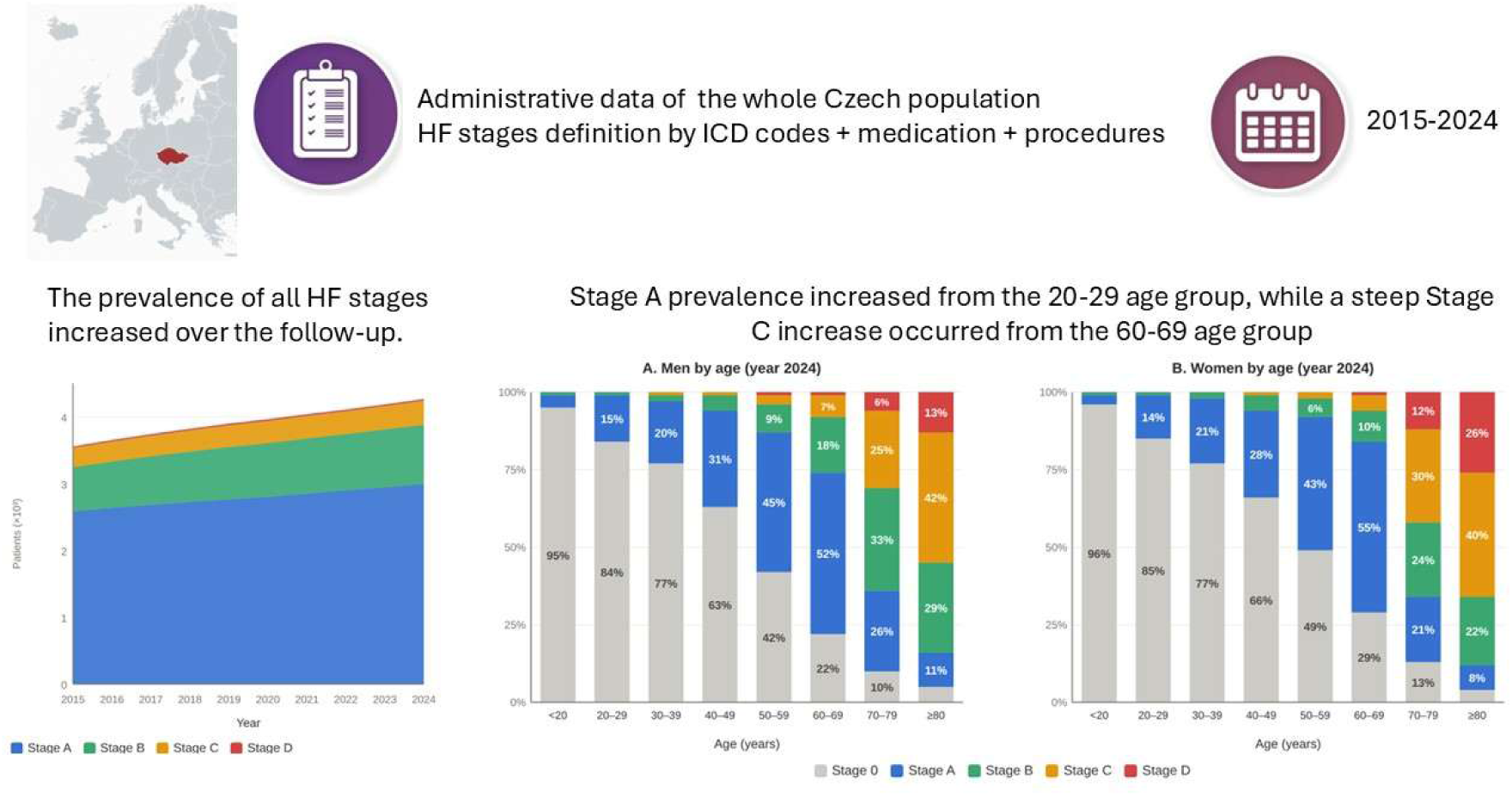

**Conclusion:** A population-wide HF staging system based on administrative data enables surveillance of the HF epidemic. In Czechia, the burden of preclinical and clinical HF is increasing.

## Introduction

Heart failure (HF) is a major and growing global public health challenge, affecting more than 64 million people worldwide and accounting for a substantial burden of disability, hospitalization, and healthcare expenditure.^1^ Although age-standardized HF incidence has stabilized or declined in many high-income countries, the overall prevalence continues to increase owing to improved survival and population aging.^2, 3^ At the same time, rising rates of obesity, diabetes, and other cardiometabolic disorders are contributing to an increasing incidence of HF among younger adults, suggesting that the future burden of HF may continue to grow despite advances in treatment.^4–6^

Heart failure develops gradually along a continuum that begins with exposure to cardiovascular risk factors (Stage A), progresses through asymptomatic structural or functional cardiac abnormalities (Stage B), and ultimately culminates in symptomatic (Stage C) and advanced HF (Stage D).^7^ Because this process often unfolds over many years, identifying individuals in preclinical stages offers an opportunity to prevent or delay overt HF through aggressive risk-factor modification and early intervention.^8–10^

Despite the importance of preclinical disease, contemporary HF epidemiology remains focused largely on symptomatic HF. Most national registries and administrative databases capture only patients with established clinical HF, whereas Stages A and B are poorly characterized at the population level. As a result, healthcare systems have limited ability to quantify the reservoir of individuals at risk for future HF, evaluate trends across the full disease continuum, or monitor the potential impact of preventive strategies. In addition, reliance on diagnostic coding alone may underestimate the burden of clinical HF, because many patients receive HF-directed pharmacotherapy without a formal HF diagnosis being recorded.

The 2021 Universal Definition and Classification of Heart Failure provides a contemporary framework for staging HF across its full clinical spectrum.^7^ However, its implementation has largely been confined to clinical cohorts with detailed imaging and biomarker assessment, limiting its application to population-wide surveillance. Whether this framework can be operationalized for nationwide surveillance using routinely collected administrative healthcare data remains uncertain.

By integrating ICD-10 diagnoses, prescription claims, and procedural records, we approximated the HF continuum across the Czech population and used linked healthcare and mortality data to examine prevalence, temporal trends, stage transitions, mortality, and treatment patterns from 2015 through 2024. Our goal was not only to describe the epidemiology of HF, but also to establish a scalable approach for monitoring the full HF continuum at the population level.

## Methods

### Data source

The present analysis used data from the National Health Information System (NHIS), managed by the Institute of Health Information and Statistics of Czechia (IHIS CR). The NHIS is established under § 70 par. 1 of Act No. 372/2011 Coll., on Health Services and Conditions of Their Provision (Act on Health Services). Due to this legal mandate, the retrospective analyses did not require either ethics committee approval or participant informed consent. For this study, data were extracted from the National Registry of Reimbursed Health Services (NRRHS), which provides an overview of insured individuals and their records from the public health insurance system. The dataset is further linked to the national death registry. Health insurance is mandatory for all residents and employees of companies based in Czechia; therefore, this population-based analysis includes all Czech citizens (10.9 million as of December 31, 2024).

### Heart failure stages classification

A staging framework for HF was introduced in 2005 based on expert consensus.^11^ Since its introduction, multiple observational and cohort studies have validated the clinical utility and prognostic relevance of this staging system by demonstrating a consistent gradient in both mortality risk and the likelihood of progression to overt HF across the stages.^12–14^ In 2021, the Universal Definition of HF revised its staging system to incorporate cardiac biomarkers.^7^ This revision increased the number of individuals classified as stage B HF.^15^

In the present study, we developed a claims-based staging framework adapted from the 2021 Universal Definition of Heart Failure using routinely collected administrative healthcare data. Because imaging findings, natriuretic peptides, and standardized symptom assessment were not available, direct implementation of the Universal Definition was not possible. We therefore introduced pragmatic modifications by integrating International Classification of Diseases (ICD-10) codes, prescription claims, and procedural records to define each HF stage. Beyond the criteria outlined in the Universal Definition, we also included selected comorbid conditions with established associations with increased HF risk. HF stages were assigned hierarchically in the following order: Stage D, Stage C, Stage B, Stage A, and Stage 0, such that each individual could belong to only one stage in a given calendar year.

**Stage A** was defined by exposure to HF risk factors—arterial hypertension, diabetes mellitus, obesity, alcohol abuse, chronic kidney disease (CKD), atherosclerotic cardiovascular disease (prior percutaneous coronary intervention or coronary artery bypass grafting without myocardial infarction history, and stroke history) and cardiotoxic cancer therapies, without any features of stage B, C, or D HF. Although CKD is not explicitly listed in the Universal Definition, we included it in stage A based on multiple lines of evidence showing that CKD is an independent risk factor for HF.^16–18^

**Stage B** was defined by evidence of structural cardiac abnormalities in the absence of criteria for Stage C or D, in accordance with the 2021 Universal Definition. We also included atrial fibrillation in Stage B, as it is associated with structural heart disease, being both a consequence of, and a contributor to, left atrial enlargement.^19^ Atrial fibrillation also independently increases the risk of developing HF.^20^ We further classified patients with pacemaker implantation under Stage B, as pacing-induced cardiomyopathy has been reported in 10–20% of individuals within 3 to 4 years post-implantation.^21^ Moreover, up to 21% of patients with right ventricular pacing develop new-onset HF.^22, 23^ In addition, we included implantable cardioverter-defibrillator (ICD) implantation in the absence of pharmacological treatment for symptomatic HF under stage B, as the most common indication for ICDs is underlying systolic dysfunction. We also classified patients receiving furosemide, spironolactone, or eplerenone without an HF-related ICD-10 code as Stage B, provided that no Stage C or D criteria were present. This reflects the fact that many patients receive HF-targeted pharmacotherapy without formal HF coding, particularly in outpatient settings, consistent with Glasgow health record data showing loop diuretic use is more strongly associated with mortality than a recorded HF diagnosis alone.^24^

To evaluate the potential influence of these pragmatic Stage B extensions, we performed a sensitivity analysis excluding individuals classified as Stage B solely on the basis of atrial fibrillation or pacemaker implantation. Under this restricted Stage B definition, the number of individuals classified as Stage B in 2024 decreased from 886,952 to 628,875, a 29.1% reduction. Despite this reduction, mortality estimates remained similar: crude 1-year mortality increased from 4.0% to 4.1%, and age-adjusted mortality increased from 1.7% to 1.9%. These findings suggest that inclusion of atrial fibrillation and pacemaker implantation increased the sensitivity of Stage B ascertainment without materially weakening the observed mortality gradient across HF stages.

**Stage C** Patients were classified as Stage C if they had: (1) an HF-specific ICD-10 code (I11.0, I13.0, I13.2, I25.5, I42.0, I42.9, I50, or R57.0) as a primary inpatient diagnosis; (2) a broader cardiac disease ICD-10 code (I05–I09, I11.0, I13.0, I13.2, I21–I25, I34–I37, I39, I40–I43, I50, Q20–Q24, or R57.0) as a secondary inpatient diagnosis or a primary outpatient specialist diagnosis, combined with a prescription for furosemide, spironolactone, eplerenone, or sacubitril-valsartan within 90 days; (3) an HF-related implantation procedure or device; or (4) a sacubitril-valsartan prescription alone, given its exclusive HF indication in the Czech reimbursement setting.

**Stage D** was defined by the history of heart transplantation, left ventricular assist device implantation, or two or more hospital admissions for HF over the past year.

Incident clinical HF in 2024 was defined as the first classification into Stage C or D in 2024 among individuals without Stage C/D classification during the preceding 10-year look-back period.

### Statistical analysis

Standard descriptive statistics were used in the analysis: absolute and relative frequencies for categorical variables and means with standard deviations for continuous variables. Incidence, prevalence, and mortality rates were age-standardized to the 2013 European Standard Population (ESP 2013), with results reported per 100,000 population. Data preprocessing was performed using the Vertica Analytic Database, followed by statistical analyses in R (R Core Team, 2025).

## Results

### Prevalence of HF stages (2015-2024)

The prevalence of all HF stages increased over the study period (**Supplementary Figure 1**). **Table 1** presents the demographic characteristics of HF stages in 2024.

**Table 1.** Characteristics of patients by heart failure stage in 2024.

|  | **Stage 0** | **Stage A** | **Stage B** | **Stage C** | **Stage D** |
| --- | --- | --- | --- | --- | --- |
|  | N = 6,516,385 | N = 2,994,456 | N = 886,952 | N = 360,094 | N = 26,474 |
| **Demographic characteristics** |  |  |  |  |  |
| Population proportion | 60.4 % | 27.8 % | 8.2 % | 3.3 % | 0.2 % |
| Male sex | 49.3 % | 48.4 % | 49.2 % | 50.4 % | 59.1 % |
| Age, mean ± SD | 32 ± 20 | 56 ± 17 | 66 ± 17 | 76 ± 13 | 75 ± 12 |
| **Medical history** |  |  |  |  |  |
| Arterial hypertension | - | 49.0 % | 62.3 % | 77.3 % | 79.7 % |
| PCI/CABG | - | 0.0 % | 11.8 % | 22.3 % | 24.7 % |
| Atrial fibrillation | - | - | 26.6 % | 43.5 % | 63.9 % |
| Stroke | - | 1.8 % | 5.3 % | 8.2 % | 9.2 % |
| Diabetes mellitus | - | 20.9 % | 30.4 % | 52.4 % | 74.0 % |
| CKD | - | 3.6 % | 8.4 % | 22.3 % | 43.2 % |
| Gastroduodenal ulcer disease | 14.6 % | 34.6 % | 52.6 % | 66.2 % | 74.9 % |
| Asthma/COPD | 16.5 % | 21.0 % | 27.3 % | 36.1 % | 44.5 % |
| Cancer in the last 5 years | 0.9 % | 4.7 % | 8.5 % | 10.4 % | 10.0 % |
| **Medication in 2024** |  |  |  |  |  |
| Digoxin (ATC C01AA05) | 0.0 % | 0.0 % | 1.4 % | 6.7 % | 13.0 % |
| Furosemide (ATC C03CA01) | - | - | 12.0 % | 58.3 % | 79.1 % |
| Spirolactone/eplerenone (ATC C03DA01, C03DA04) | - | - | 8.5 % | 39.8 % | 62.0 % |
| Other diuretics (ATC C03 without C03CA01, C03DA01, C03DA04) | 0.0 % | 6.2 % | 10.3 % | 8.4 % | 4.4 % |
| Beta-blockers (ATC C07) | 0.2 % | 19.7 % | 45.3 % | 68.5 % | 80.1 % |
| Calcium channel blockers (ATC C08) | 0.1 % | 11.1 % | 18.1 % | 21.8 % | 14.9 % |
| ARNI (ATC C09DX04) | - | - | - | 5.6 % | 24.3 % |
| ACE inhibitors/ARBs (ATC C09 without C09DX04) | 0.5 % | 53.9 % | 61.4 % | 66.8 % | 49.7 % |
| Dapagliflozin/empagliflozin (ATC A10BK01, A10BK03) | 0.0 % | 2.3 % | 5.9 % | 23.5 % | 50.8 % |

CABG – coronary artery bypass graft, CKD – chronic kidney disease, COPD – chronic obstructive pulmonary disease, PCI – percutaneous coronary intervention, ARNI – angiotensin receptor/neprilysin inhibitor, ACEi – Angiotensin-converting enzyme inhibitors, AT2 – Angiotensin II receptor inhibitors

#### Stage A

The crude prevalence of individuals with Stage A HF increased from 247.7 per 1,000 in 2015 to 277.6 per 1,000 in 2024, a 12.1% rise (**Table 2**). Additionally, the age-standardized prevalence increased from 244.1 per 1,000 in 2015 to 267.2 per 1,000 in 2024, a 9.46% increase.

**Table 2.**
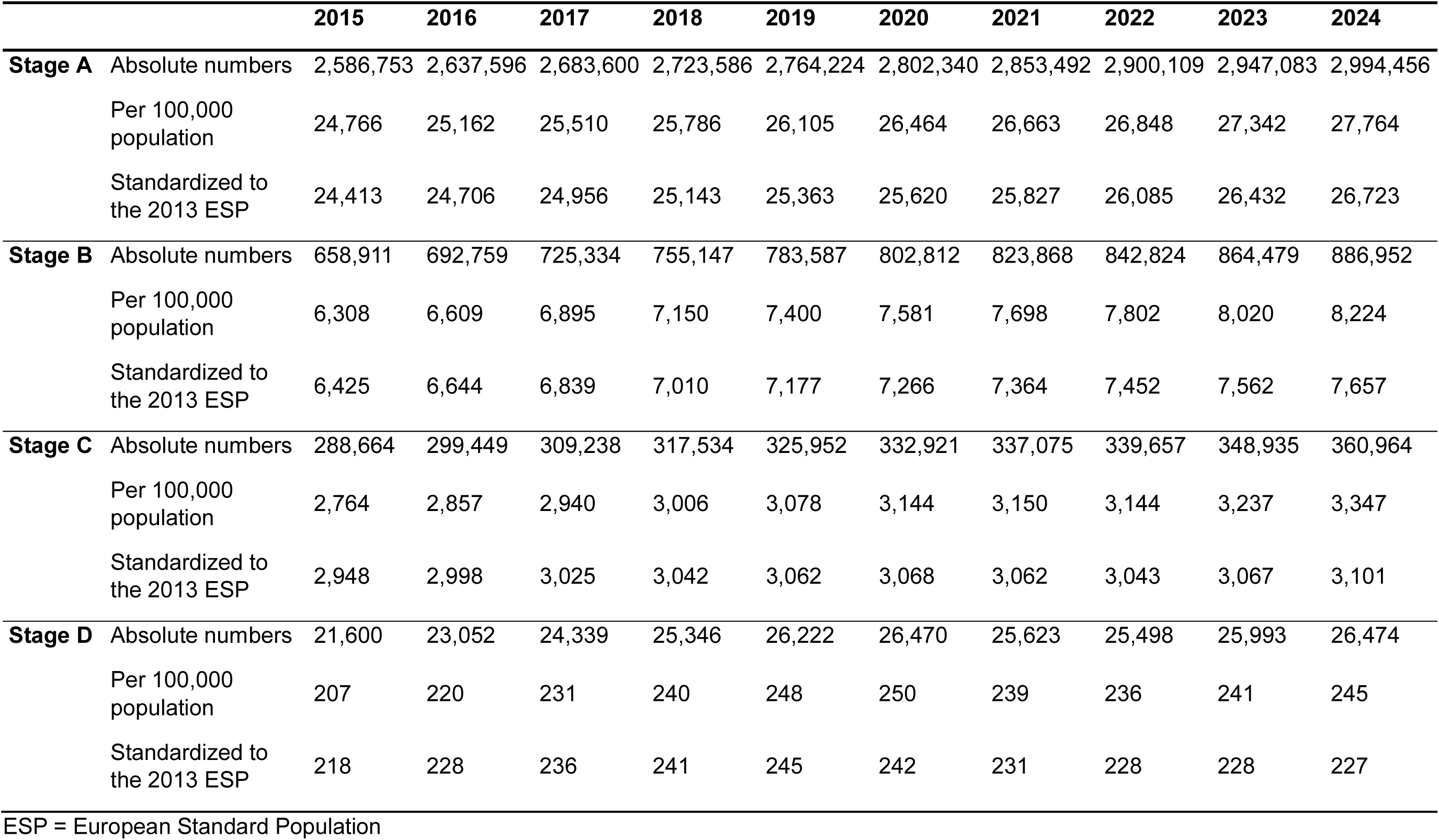
Prevalence and time trends of heart failure stages.

|  |  | 2015 | 2016 | 2017 | 2018 | 2019 | 2020 | 2021 | 2022 | 2023 | 2024 |
| --- | --- | --- | --- | --- | --- | --- | --- | --- | --- | --- | --- |
| **Stage A** | Absolute numbers | 2,586,753 | 2,637,596 | 2,683,600 | 2,723,586 | 2,764,224 | 2,802,340 | 2,853,492 | 2,900,109 | 2,947,083 | 2,994,456 |
|  | Per 100,000 population | 24,766 | 25,162 | 25,510 | 25,786 | 26,105 | 26,464 | 26,663 | 26,848 | 27,342 | 27,764 |
|  | Standardized to the 2013 ESP | 24,413 | 24,706 | 24,956 | 25,143 | 25,363 | 25,620 | 25,827 | 26,085 | 26,432 | 26,723 |
| **Stage B** | Absolute numbers | 658,911 | 692,759 | 725,334 | 755,147 | 783,587 | 802,812 | 823,868 | 842,824 | 864,479 | 886,952 |
|  | Per 100,000 population | 6,308 | 6,609 | 6,895 | 7,150 | 7,400 | 7,581 | 7,698 | 7,802 | 8,020 | 8,224 |
|  | Standardized to the 2013 ESP | 6,425 | 6,644 | 6,839 | 7,010 | 7,177 | 7,266 | 7,364 | 7,452 | 7,562 | 7,657 |
| **Stage C** | Absolute numbers | 288,664 | 299,449 | 309,238 | 317,534 | 325,952 | 332,921 | 337,075 | 339,657 | 348,935 | 360,964 |
|  | Per 100,000 population | 2,764 | 2,857 | 2,940 | 3,006 | 3,078 | 3,144 | 3,150 | 3,144 | 3,237 | 3,347 |
|  | Standardized to the 2013 ESP | 2,948 | 2,998 | 3,025 | 3,042 | 3,062 | 3,068 | 3,062 | 3,043 | 3,067 | 3,101 |
| **Stage D** | Absolute numbers | 21,600 | 23,052 | 24,339 | 25,346 | 26,222 | 26,470 | 25,623 | 25,498 | 25,993 | 26,474 |
|  | Per 100,000 population | 207 | 220 | 231 | 240 | 248 | 250 | 239 | 236 | 241 | 245 |
|  | Standardized to the 2013 ESP | 218 | 228 | 236 | 241 | 245 | 242 | 231 | 228 | 228 | 227 |

ESP = European Standard Population

#### Stage B

The crude prevalence of Stage B increased from 63.1 per 1,000 in 2015 to 82.2 per 1,000 in 2024, a 30.4% rise. The age-standardized prevalence also rose by 19.2%, from 64 per 1,000 to 77 per 1,000.

#### Stages C and D

The crude prevalence of Stage C increased from 28 per 1,000 in 2015 to 33 per 1,000 in 2024, reflecting a 21.1% rise. The age-standardized prevalence also rose by 5.2%, from 29.5 per 1,000 to 31.0 per 1,000. After 2020, however, the standardized prevalence of Stage C increased only modestly.

Similarly, between 2015 and 2024, the crude prevalence of Stage D increased by 18.4%, while the age-standardized prevalence rose by 4.1%. However, a nonlinear trend in standardized prevalence was observed, with a peak in 2019 followed by a plateau, coinciding with the COVID-19 period and possibly reflecting increased mortality in this high-risk group (**Supplementary Figure 2**).

#### Age-specific distribution of HF stages

To determine the age at which the prevalence of each HF stage begins to increase, we plotted the 2024 stage-specific prevalence across the age groups (**Figure 1**). The proportion of individuals classified as Stage A increased from early adulthood through the 60–69 age group and then declined at older ages as Stages B–D became increasingly prevalent. No difference in pattern was observed between men and women. Stage B exhibited a sharp increase in prevalence starting from the 40–49 age group. The prevalence of Stage C began to rise from the 60–69 age group onward, with a similar pattern of increase in women and men.

**Figure 1:**
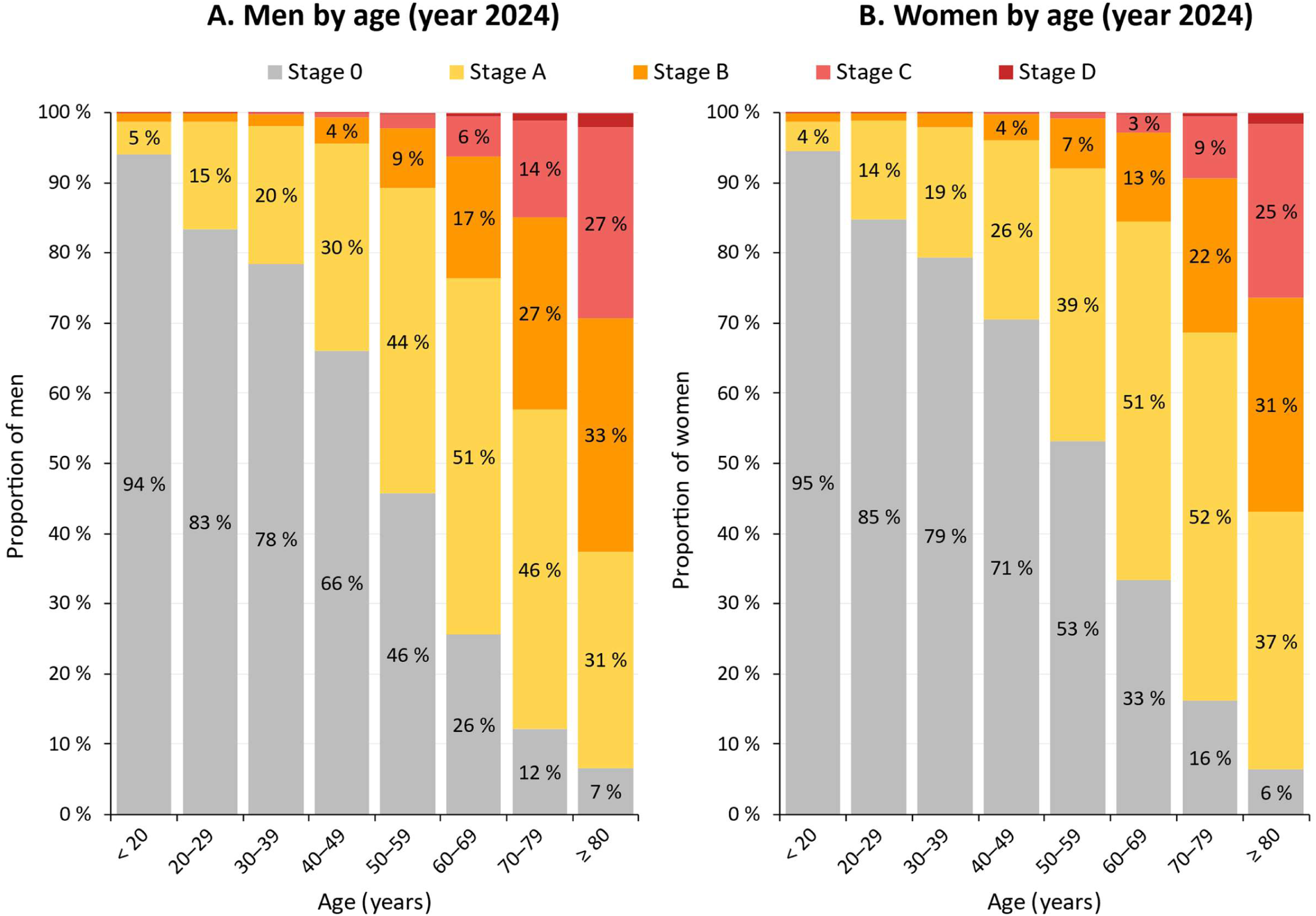
Distribution of heart failure stages in 2024 by age and sex

### Transition of HF stages

Among individuals classified as Stage A in 2015, 13.2% were classified as Stage B in 2024, 4.9% as Stage C, and 0.3% as Stage D by 2024 (**Figure 2, Supplementary table 3**). In 2024, among 52,172 individuals with incident clinical HF, more than 95% had previously been classified as Stage A or B.

**Figure 2:**
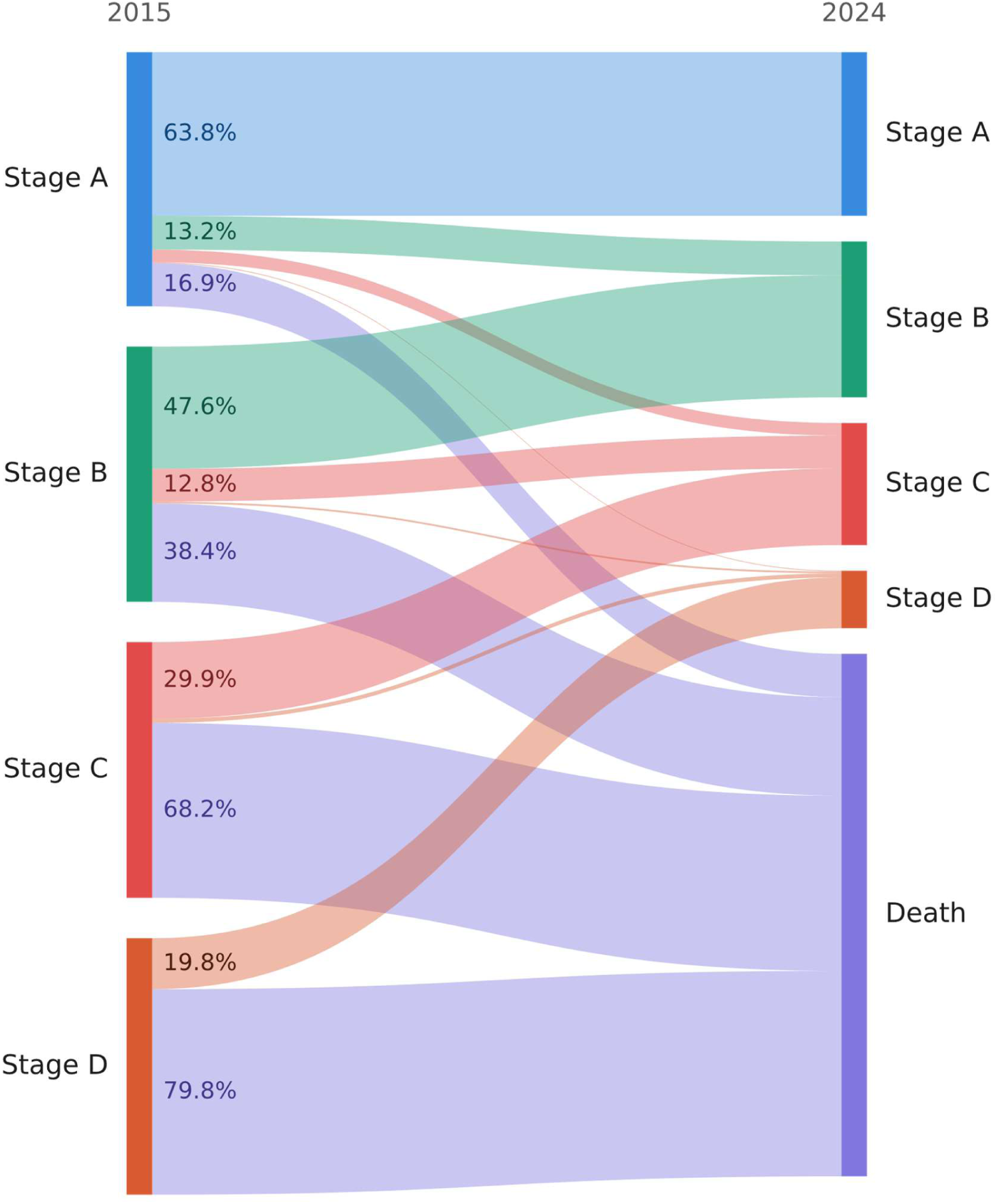
Transition of heart failure stages during the follow-up (2015-2024)

### Mortality risk by HF stages

Analysis of data from 2024 demonstrated a stepwise rise in 1-year mortality with advancing heart failure stage (**Figure 3**). Unstandardized and age-standardized 1-year mortality rates were 0.12% and 0.45% for Stage 0, rising to 0.97% and 0.69% in Stage A, 4.00% and 1.69% in Stage B, 9.49% and 3.06% in Stage C, and 18.73% and 7.27% in Stage D, respectively (**Table 3**). Compared with Stage 0, the age-standardized relative mortality risk rose progressively across HF stages—1.53 in Stage A, 3.76 in Stage B, 6.8 in Stage C, and 16.2 in Stage D. As the HF stage increased, the proportion of deaths attributed to cardiovascular causes rose from 21.5% in Stage 0 to 60.2% in Stage D, while the proportion of deaths due to neoplasms declined from 34.5% in Stage 0 to 9.4% in Stage D (**Supplementary Table 2**).

**Figure 3:**
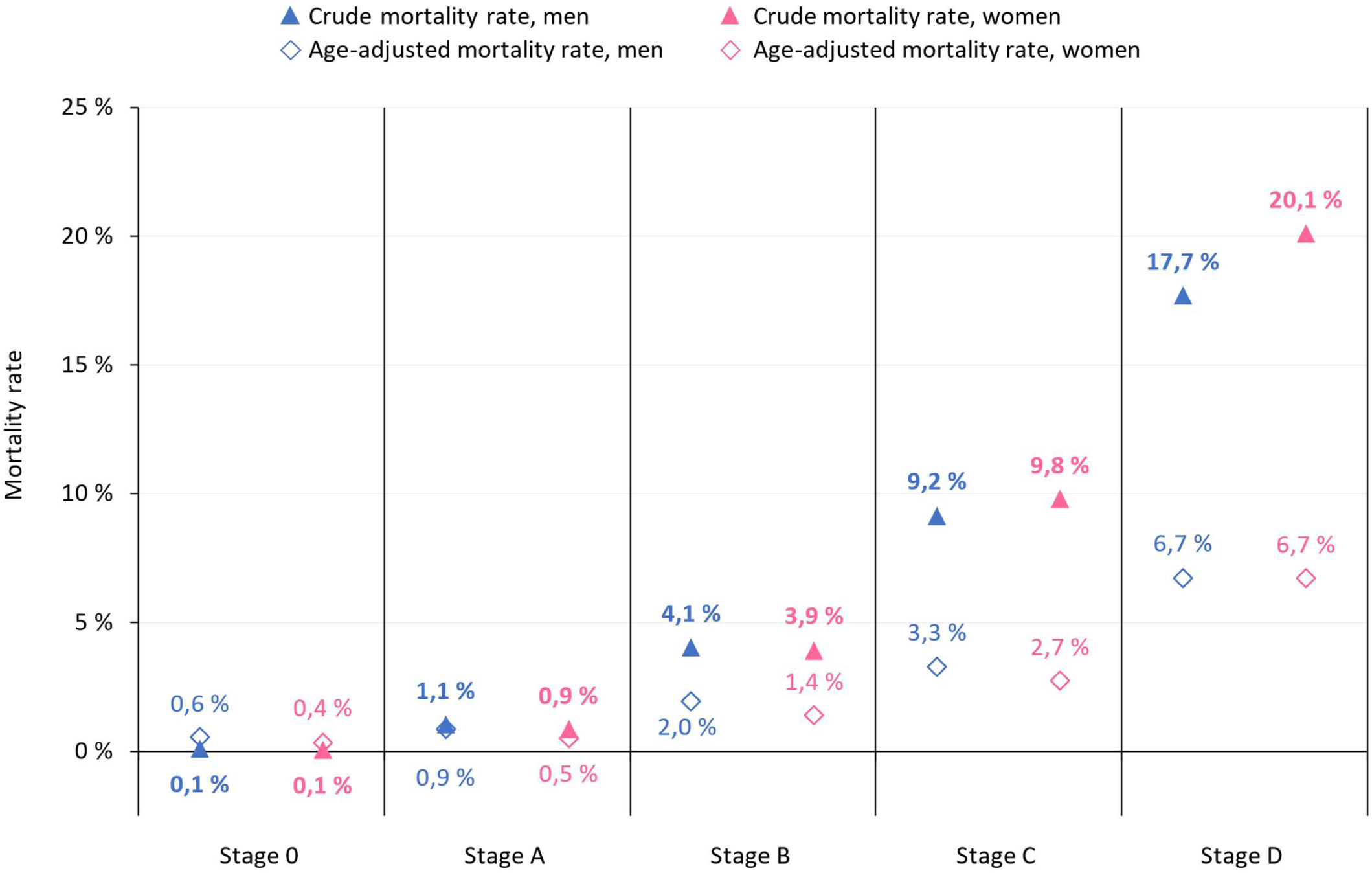
Mortality rate by heart failure stages in 2024

**Figure 4:**
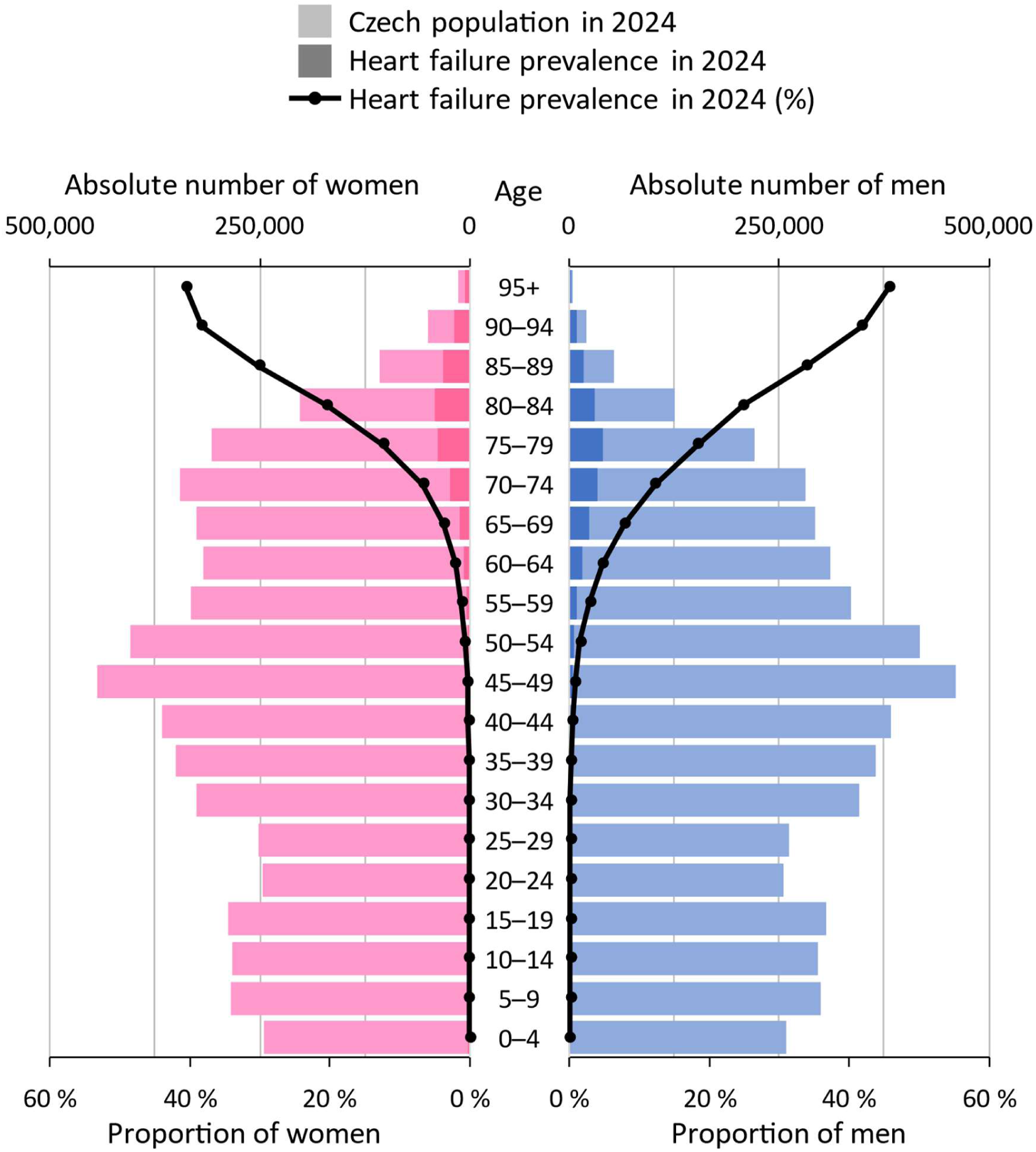
Age- and sex-specific distribution of clinical heart failure (Stages C and D) in the Czech population in 2024.

**Table 3.** Annual mortality by the heart failure stages.

|  | 2015 | 2016 | 2017 | 2018 | 2019 | 2020 | 2021 | 2022 | 2023 | 2024 |
| --- | --- | --- | --- | --- | --- | --- | --- | --- | --- | --- |
| **Stage 0** Annual mortality | 0.15 % | 0.14 % | 0.14 % | 0.14 % | 0.14 % | 0.14 % | 0.16 % | 0.14 % | 0.12 % | 0.12 % |
| Standardized to the 2013 ESP | 0.62 % | 0.58 % | 0.59 % | 0.55 % | 0.53 % | 0.60 % | 0.64 % | 0.55 % | 0.47 % | 0.45 % |
| **Stage A** Annual mortality | 1.09 % | 1.04 % | 1.05 % | 1.07 % | 1.04 % | 1.18 % | 1.33 % | 1.09 % | 0.99 % | 0.97 % |
| Standardized to the 2013 ESP | 0.80 % | 0.75 % | 0.77 % | 0.78 % | 0.77 % | 0.83 % | 0.93 % | 0.79 % | 0.72 % | 0.69 % |
| **Stage B** Annual mortality | 5.21 % | 4.76 % | 4.74 % | 4.61 % | 4.43 % | 5.02 % | 5.28 % | 4.47 % | 4.11 % | 4.00 % |
| Standardized to the 2013 ESP | 2.27 % | 2.03 % | 2.03 % | 1.99 % | 1.90 % | 2.06 % | 2.24 % | 1.94 % | 1.74 % | 1.69 % |
| **Stage C** Annual mortality | 11.00 % | 10.41 % | 10.60 % | 10.51 % | 10.35 % | 11.92 % | 12.31 % | 10.69 % | 9.79 % | 9.49 % |
| Standardized to the 2013 ESP | 3.81 % | 3.53 % | 3.93 % | 3.49 % | 3.51 % | 3.83 % | 4.16 % | 3.42 % | 3.15 % | 3.06 % |
| **Stage D** Annual mortality | 23.24 % | 22.17 % | 21.78 % | 22.09 % | 21.34 % | 23.64 % | 22.66 % | 20.80 % | 19.45 % | 18.73 % |
| Standardized to the 2013 ESP | 8.54 % | 11.88 % | 9.38 % | 9.23 % | 9.19 % | 9.74 % | 8.48 % | 8.29 % | 7.89 % | 7.27 % |

ESP = European Standard Population

### Pharmacotherapy of symptomatic HF stages

A substantial treatment gap was observed in the pharmacotherapy of symptomatic HF stages. Among patients in Stage C, 24% received sodium-glucose cotransporter 2 (SGLT2) inhibitors. This has increased by more than half compared with 2023. Just 6% received angiotensin receptor–neprilysin inhibitors (ARNIs). Although these rates remain suboptimal, they should be interpreted in the context of the rapid implementation of SGLT2 inhibitors in the Czech Republic. Reimbursement for dapagliflozin in heart failure with reduced ejection fraction (HFrEF) became available in February 2022, followed by reimbursement for empagliflozin across the full spectrum of left ventricular ejection fraction in May 2023 and for dapagliflozin in March 2024. In Stage D, the use of guideline-recommended therapies increased modestly, with 51% receiving SGLT2 inhibitors and 24% receiving ARNIs. In contrast, 15% of patients in Stage D were prescribed calcium channel blockers, a therapy that is generally not part of guideline-directed treatment for advanced HFrEF and may reflect competing indications or residual prescribing inertia (**Table 1**).

## Discussion

In this nationwide study of 10.9 million Czech residents, we developed and applied a nationwide claims-based staging framework aligned with the 2021 Universal Definition of HF, using routinely collected administrative healthcare data to approximate the HF continuum over a 10-year period. Four principal findings emerge. First, we demonstrate that nationwide administrative data can be used to classify all HF stages, enabling surveillance of the disease continuum beyond clinically overt HF. Second, the prevalence of both preclinical HF stages increased substantially over time, even after age standardization, indicating that the growing HF burden cannot be explained by population aging alone. Third, more than 95% of individuals who developed clinical HF had previously been classified as Stage A or B, highlighting a prolonged and potentially modifiable preclinical phase. Finally, despite recent improvements, substantial gaps remain in the implementation of guideline-directed medical therapy among patients with symptomatic HF.

Current epidemiological surveillance primarily focuses on symptomatic HF, whereas the preclinical stages remain poorly characterized at the population level. Most available data originate from prospective cohort studies,^12, 25–27^ which provide valuable mechanistic insights but are limited by selective recruitment, restricted age ranges, and relatively small sample sizes. Consequently, they are not ideally suited for monitoring nationwide trends or informing healthcare planning. By contrast, administrative healthcare databases encompass entire populations and are continuously updated, making them an attractive resource for population-level surveillance. Until now, however, the absence of a standardized administrative definition of HF stages has limited their application to the full HF continuum.

To address this gap, we operationalized the 2021 Universal Definition of HF within routinely collected administrative healthcare data. Because administrative datasets lack imaging and biomarker information required by the original definition, our framework incorporates pragmatic modifications based on diagnostic codes, prescription claims, and procedural records.

We also refined the definition of symptomatic HF to improve case ascertainment. Because HF is frequently undercoded in administrative databases, particularly among patients with preserved ejection fraction or those managed without hospitalization, we supplemented HF-specific diagnostic codes with a combination of broader cardiac disease codes and pharmacotherapy strongly suggestive of symptomatic HF (furosemide, spironolactone, eplerenone, or sacubitril-valsartan) within 90 days of the diagnostic code, as an indicator of Stage C disease. A prescription for sacubitril-valsartan alone was also sufficient to classify a patient as Stage C, given its near-exclusive indication for symptomatic HF.

These adaptations reflect the inherent limitations of administrative data, which do not capture echocardiographic findings or cardiac biomarkers required by the Universal Definition. Although this approach cannot achieve the diagnostic precision of comprehensive clinical phenotyping, it enables standardized HF staging at the population level using routinely available healthcare data. Future validation against echocardiographic registries and clinical datasets incorporating imaging and biomarker information will be essential to further establish the diagnostic performance of this administrative staging algorithm. The resulting framework demonstrated strong construct validity, with a progressive increase in both all-cause and cardiovascular mortality across successive HF stages, closely mirroring observations from community-based cohort studies.^12, 15^ Together, these findings support the use of this framework for nationwide surveillance of the HF continuum and for facilitating comparisons of HF burden across healthcare systems.

A particularly important finding is the substantial increase in the prevalence of Stage A and Stage B HF during the study period. Although population aging contributes to the increasing burden of HF, age-standardized prevalence also rose considerably, indicating that demographic changes alone do not explain these trends. While causal inferences cannot be drawn from administrative data, the increasing prevalence of obesity and cardiometabolic disease in Czechia is a plausible contributor.^28^ Rising obesity rates not only increase the risk of HF directly but also promote hypertension, diabetes mellitus, chronic kidney disease, and atrial fibrillation, thereby accelerating progression along the HF continuum. These findings suggest that the future burden of symptomatic HF will depend increasingly on the effectiveness of primary prevention strategies targeting cardiometabolic risk factors.

Perhaps the most clinically relevant observation is that more than 95% of individuals with incident clinical HF had previously fulfilled administrative criteria for Stage A or Stage B. Although this proportion depends on the performance of the staging algorithm, it demonstrates that progression to overt HF is rarely abrupt and is typically preceded by years of identifiable cardiovascular risk or structural heart disease. This prolonged preclinical phase may represent the greatest opportunity for prevention. Our findings complement evidence from cohort studies suggesting that the majority of incident HF is attributable to a relatively small number of modifiable cardiovascular risk factors^29^ and are consistent with recent recommendations advocating risk-based primary prevention of HF. Together, these observations support a shift in healthcare policy from reactive management of symptomatic HF toward earlier identification and intervention among individuals at increased risk, as outlined by the 2025 AHA Scientific Statement on risk-based primary prevention of HF.^30^

Our analysis also provides a contemporary overview of HF pharmacotherapy at the national level. Uptake of sodium-glucose cotransporter-2 inhibitors increased substantially between 2023 and 2024, reflecting the rapid implementation of recent guideline recommendations. Nevertheless, use of angiotensin receptor–neprilysin inhibitors remained low, and a considerable proportion of patients with advanced HF continued to receive medications with limited or uncertain benefit. Because all major guideline-directed HF therapies are reimbursed within the Czech healthcare system, these findings suggest that barriers to implementation are more likely related to therapeutic inertia, clinician awareness, or healthcare organization than to treatment availability.

The prevalence of Stage A and Stage B observed in our study was lower than that reported in community cohorts incorporating systematic echocardiography and biomarker assessment.^14, 31^ This difference is expected because administrative databases cannot identify subclinical structural abnormalities, diastolic dysfunction, or elevated natriuretic peptide concentrations in asymptomatic individuals. Consequently, our estimates of preclinical HF should be regarded as conservative lower bounds. Conversely, the prevalence of symptomatic HF exceeded that reported in several Western European and North American populations,^32, 33^ suggesting that the burden of clinical HF in Czechia remains substantial and deserves continued public health attention.

Beyond the findings from Czechia, the proposed administrative staging framework may provide a standardized methodology for monitoring the HF continuum across healthcare systems. This framework could serve as a standardized methodology for international comparisons of HF epidemiology using routinely collected healthcare data, thereby facilitating benchmarking of disease burden, preventive strategies, and healthcare delivery.

### Limitations

This study has several limitations. Administrative healthcare data are collected primarily for reimbursement rather than research purposes and therefore inevitably introduce some degree of disease misclassification. Several Stage A risk factors, including obesity and alcohol misuse, are incompletely captured by diagnostic coding, while important Stage B characteristics such as ventricular remodeling, diastolic dysfunction, and elevated cardiac biomarkers are unavailable. The proposed staging framework therefore cannot replace comprehensive clinical assessment. Furthermore, individual-patient-level validation against echocardiographic registries and datasets containing imaging and biomarker data will be necessary to quantify the sensitivity, specificity, and predictive value of the proposed staging algorithm. We cannot rule out the possibility that observed secular trends in stage prevalence may partly reflect changes in coding practices, diagnostic intensity, reimbursement policies, and uptake of cardiovascular therapies over time, rather than true changes in disease burden alone. Finally, although our findings are based on nationwide data with complete population coverage, validation in other healthcare systems will be important to establish the generalizability of this approach. Nevertheless, the consistent stepwise increase in all-cause and cardiovascular mortality across stages supports the construct validity of the framework.

## Conclusion

In conclusion, routinely collected administrative healthcare data can be used to stage the HF continuum and support nationwide surveillance of both preclinical and clinical disease. In Czechia, the prevalence of preclinical HF increased substantially over time beyond what would be expected from population aging alone, indicating a growing reservoir of individuals at risk for future symptomatic HF. More broadly, this work provides a scalable framework for monitoring HF epidemiology, evaluating preventive strategies, and benchmarking HF burden across healthcare systems.

## Disclaimer

None

## Data sharing statement

Individual participant data that underlie the results reported in this article will be available to researchers who provide a methodologically sound proposal with pre-defined aims.

---

**Supplementary Table 1:**
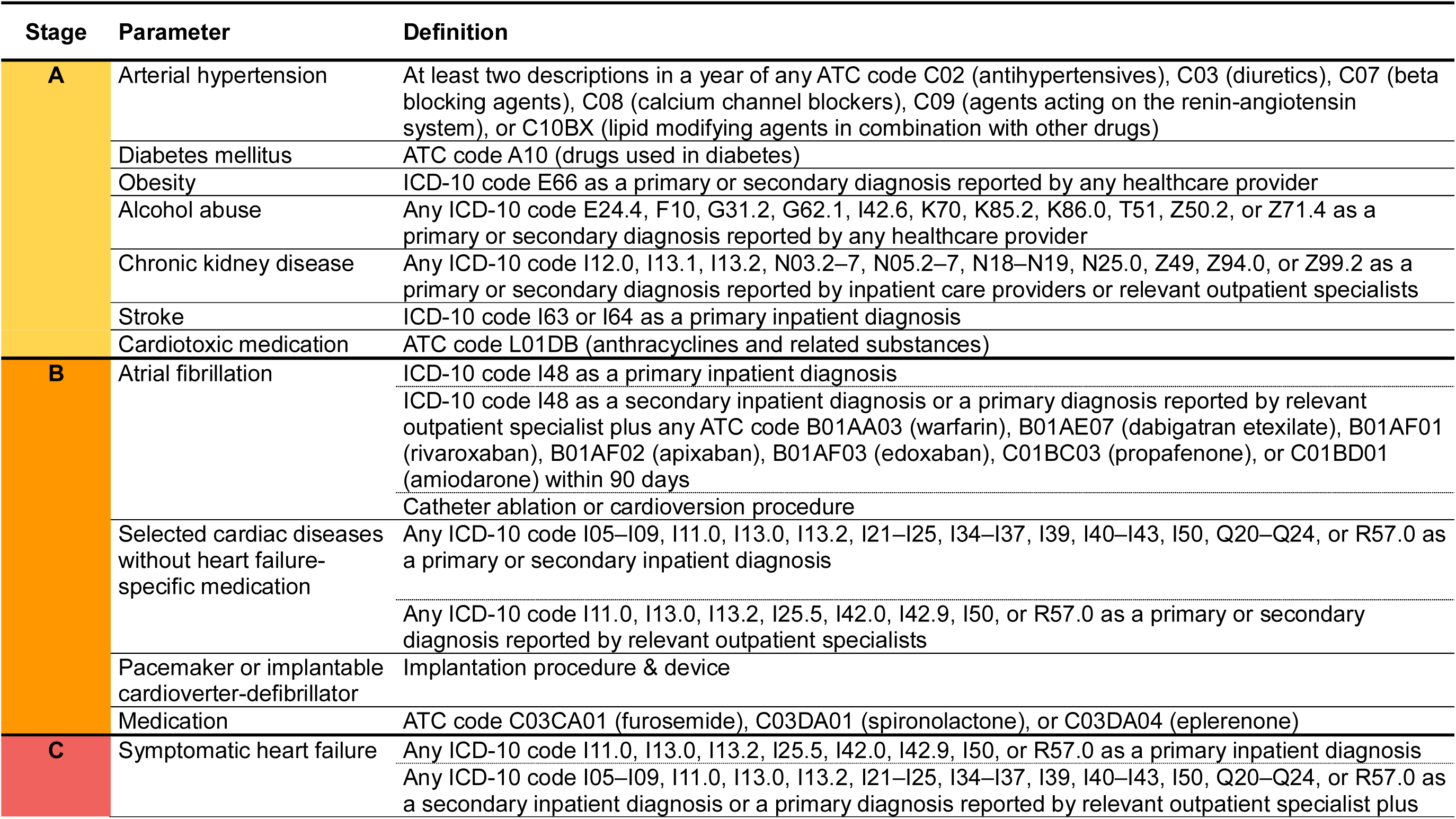
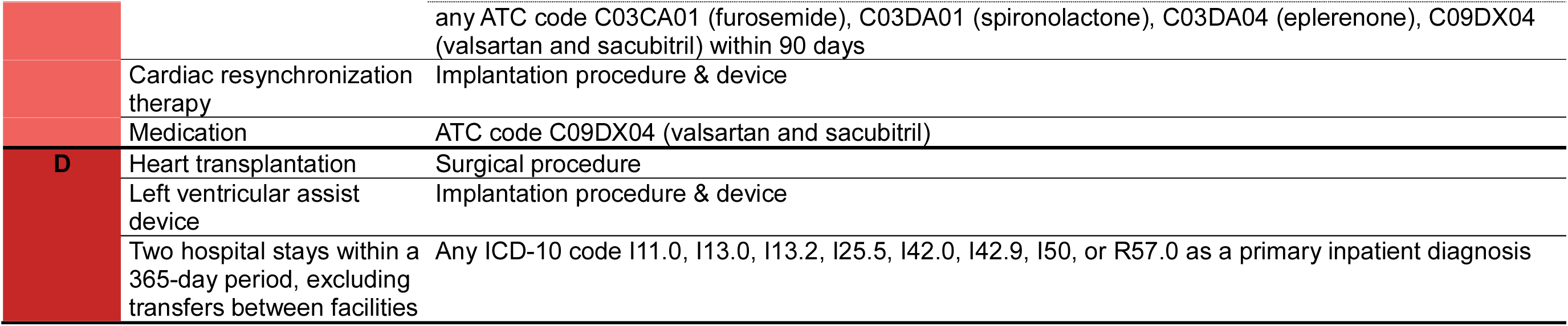
Definition of heart failure stages.

**Supplementary Table 2:** Causes of death by heart failure stage among patients deceased in 2024.

| **Chapters of ICD-10: causes of death** | **Stage 0** | **Stage A** | **Stage B** | **Stage C** | **Stage D** |
| --- | --- | --- | --- | --- | --- |
| Number of death | 7,759 | 28,971 | 35,482 | 34,259 | 4,958 |
| **I** Certain infectious and parasitic diseases | 1.5 % | 1.8 % | 2.2 % | 2.7 % | 2.5 % |
| **II** Neoplasms | 34.5 % | 31.5 % | 28.1 % | 16.7 % | 9.4 % |
| **III** Diseases of the blood and blood-forming organs and certain disorders involving the immune mechanism | 0.2 % | 0.2 % | 0.3 % | 0.3 % | 0.3 % |
| **IV** Endocrine, nutritional and metabolic diseases | 1.3 % | 4.4 % | 5.0 % | 6.7 % | 6.5 % |
| **V** Mental and behavioral disorders | 1.5 % | 2.2 % | 1.8 % | 1.3 % | 0.8 % |
| **VI** Diseases of the nervous system | 6.7 % | 5.4 % | 4.2 % | 2.8 % | 0.9 % |
| **IX** Diseases of the circulatory system | 21.5 % | 29.9 % | 35.8 % | 48.1 % | 60.2 % |
| **X** Diseases of the respiratory system | 6.5 % | 6.0 % | 7.7 % | 9.4 % | 9.0 % |
| **XI** Diseases of the digestive system | 3.1 % | 5.4 % | 6.2 % | 3.7 % | 2.6 % |
| **XII** Diseases of the skin and subcutaneous tissue | 0.1 % | 0.1 % | 0.3 % | 0.3 % | 0.2 % |
| **XIII** Diseases of the musculoskeletal system and connective tissue | 0.2 % | 0.2 % | 0.3 % | 0.3 % | 0.2 % |
| **XIV** Diseases of the genitourinary system | 0.7 % | 1.2 % | 1.8 % | 2.7 % | 3.2 % |
| **XVI** Certain conditions originating in the perinatal period | 1.2 % | 0.0 % | 0.0 % | 0.0 % | 0.0 % |
| **XVII** Congenital malformations, deformations and chromosomal abnormalities | 0.7 % | 0.1 % | 0.1 % | 0.1 % | 0.1 % |
| **XVIII** Symptoms, signs and abnormal clinical and laboratory findings, not elsewhere classified | 5.9 % | 3.3 % | 2.2 % | 1.9 % | 1.6 % |
| **XIX** Injury, poisoning and certain other consequences of external causes | 14.0 % | 7.7 % | 3.4 % | 2.2 % | 1.7 % |
| **XXII** Codes for special purposes | 0.4 % | 0.5 % | 0.7 % | 0.8 % | 0.7 % |

**Supplementary table 3:**
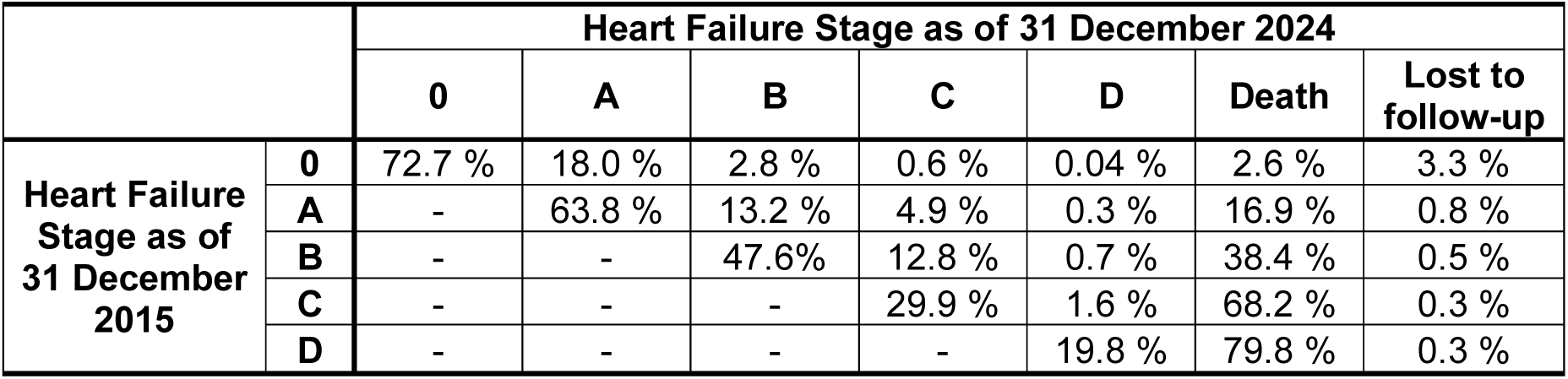
Transition of heart failure stages during the follow-up.

|  |  | Heart Failure Stage as of 31 December 2024 |  |  |  |  |  |  |
| --- | --- | --- | --- | --- | --- | --- | --- | --- |
|  |  | 0 | A | B | C | D | Death | Lost to follow-up |
| Heart Failure Stage as of 31 December 2015 | 0 | 72.7 % | 18.0 % | 2.8 % | 0.6 % | 0.04 % | 2.6 % | 3.3 % |
|  | A | - | 63.8 % | 13.2 % | 4.9 % | 0.3 % | 16.9 % | 0.8 % |
|  | B | - | - | 47.6% | 12.8 % | 0.7 % | 38.4 % | 0.5 % |
|  | C | - | - | - | 29.9 % | 1.6 % | 68.2 % | 0.3 % |
|  | D | - | - | - | - | 19.8 % | 79.8 % | 0.3 % |

**Table 4.** Epidemiology of clinical heart failure stages (stage C+D)

|  |  | 2015 | 2016 | 2017 | 2018 | 2019 | 2020 | 2021 | 2022 | 2023 | 2024 |
| --- | --- | --- | --- | --- | --- | --- | --- | --- | --- | --- | --- |
| **Incidence** | Absolute numbers | 52,171 | 49,292 | 47,599 | 47,568 | 48,534 | 46,835 | 49,462 | 50,042 | 51,796 | 52,172 |
|  | Per 100,000 population | 499.5 | 470.2 | 452.5 | 450.4 | 458.4 | 442.3 | 462.2 | 463.3 | 480.7 | 483.7 |
|  | Standardized to the 2013 ESP | 526.2 | 488.0 | 460.2 | 449.8 | 450.3 | 426.9 | 444.3 | 444.1 | 450.8 | 444.4 |
| **Prevalence** | Absolute numbers | 310,264 | 322,501 | 333,577 | 342,880 | 352,174 | 359,391 | 362,698 | 365,155 | 374,928 | 387,438 |
|  | Per 100,000 population | 2,970 | 3,077 | 3,171 | 3,246 | 3,326 | 3,394 | 3,389 | 3,380 | 3,478 | 3,592 |
|  | Standardized to the 2013 ESP | 3,166 | 3,226 | 3,261 | 3,282 | 3,307 | 3,311 | 3,293 | 3,271 | 3,295 | 3,328 |
| **Mortality** | Absolute numbers | 36,779 | 36,287 | 38,072 | 38,981 | 39,342 | 45,928 | 47,298 | 41,615 | 39,209 | 39,217 |
|  | Per 100,000 population | 352.1 | 346.2 | 361.9 | 369.1 | 371.5 | 433.7 | 442.0 | 385.2 | 363.8 | 363.6 |
|  | Standardized to the 2013 ESP | 400.1 | 384.8 | 394.0 | 393.6 | 389.2 | 443.3 | 449.1 | 394.3 | 363.5 | 353.2 |

ESP = European Standard Population

**Supplementary Figure 1:**
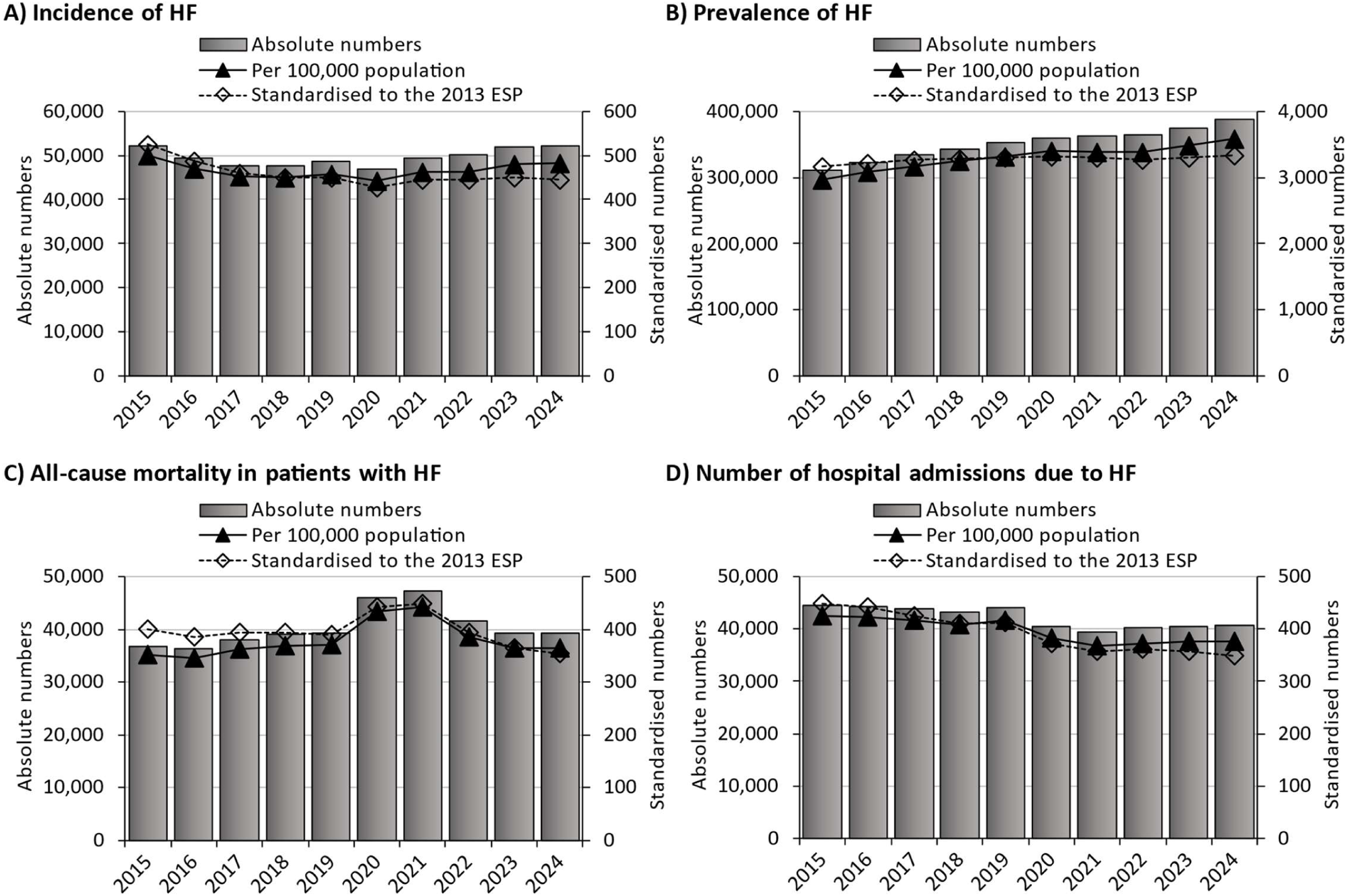
Incidence, prevalence, all-cause mortality, and hospital admissions for heart hart failure between 2015-2024.

**Supplementary Figure 2:**
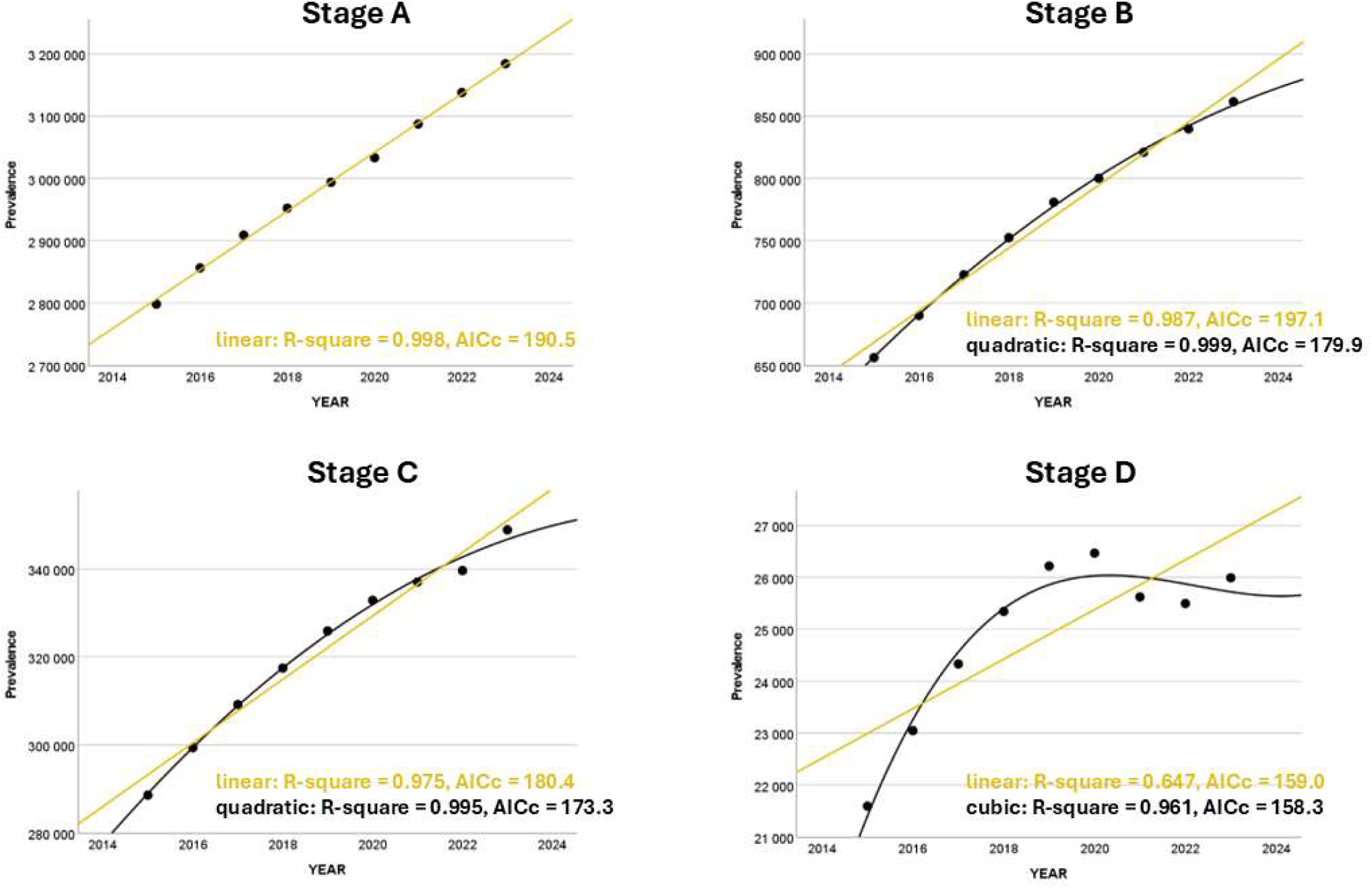
Trends in heart failure stages

